# *“I was facilitating everybody else’s life. And mine had just ground to a halt”*: the COVID-19 pandemic and its impact on women in the UK

**DOI:** 10.1101/2021.03.12.21253136

**Authors:** Asha Herten-Crabb, Clare Wenham

## Abstract

A growing body of research has highlighted the disproportionately negative impact of the COVID-19 pandemic on women globally. This paper contributes to this work by interrogating the lived realities of 64 women in the UK through semi-structured interviews, undertaken during the first and second periods of lockdown associated with COVID-19 in 2020. Categorising the data by theme and type of gendered disadvantage, this paper explores the normative and policy-imposed constraints experienced by women in 2020, highlighting the role that government can and should proactively play in attending to gender inequalities throughout its COVID-19 response.

## Introduction

*“women will just pick up the childcare, women will pick up the care of elderly parents, they will shoulder the burden of whatever policies are put in place, or decisions that are made, they will just get on with it”*

Infectious disease outbreaks infect and affect men, women and non-binary genders differently. In the ongoing COVID-19 pandemic, current data suggest that men are infected more than women with a ratio of 1.1:1, and that men suffer more severe symptoms and greater mortality (Global Health 50/50 2020). However, the downstream effects of COVID-19 are also heavily gendered and affect women disproportionately to men. In this paper, we show results from qualitative research to show the socio-economic effects of non-pharmaceutical interventions to stem the spread of the virus have fallen differentially on women in the United Kingdom (UK).

These direct and indirect gendered effects of outbreak response interventions have been documented in previous outbreaks. During Ebola in West Africa, women’s caretaking role within communities exposed them to greater risk than male counterparts, as it was women who cared for the sick, and/or were responsible for washing the bodies of the deceased prior to burial, a key mechanism for transmission (Nkangu, Olatunde, and Yaya 2017). This disproportionately exposed women to an increased risk of Ebola virus infection relative to men (WHO 2016).

Women were also disproportionately impacted economically. In Sierra Leone, markets closed for quarantine purposes, in which most of the traders were women. This placed women at risk of further socio-economic insecurity. Bandiera et al. (2019) have shown that the impact of these closures were long-lasting: 13 months after the start of the crisis 63% of men had returned to work, compared to only 17% of women. This was echoed during Zika: the sector of the economy most negatively impacted was the tourism, which predominantly employs women. These impacts were amplified at the micro-level, as it was women who had children born with Congenital Zika Syndrome that were unable to continue in employment due to the physical needs of their children (Human Rights Watch 2017).

Women also faced increased physical and structural violence because of Ebola and Zika. The Ebola outbreak in West Africa revealed a secondary epidemic of rape and sexual violence as quarantine orders were implemented (Yasmin 2016), witnessed again during the Ebola outbreak in DRC in 2019 (International Rescue Committee 2019). This direct violence was compounded by barriers to accessing support: within health systems, women would be less likely to access healthcare providers, be less able to afford time off work if sick or with sick family members, be unable to access routine water and sanitation facilities to reduce their risk of infection (anonymous, forthcoming). Thus, historically women experience outbreaks and the secondary effects of government policies to respond to outbreaks differently to men.

2020 has witnessed a number of editorials, working papers and reports warning of the likely disproportionate impact of COVID-19 on women (e.g. Hupkau and Petrongolo, 2020; Peterman *et al*., 2020; Wenham, Smith and Morgan, 2020). Despite these calls for gender-responsive interventions, primary research from the first 10 months of the pandemic shows that women’s lives around the world have been disproportionately negatively affected by COVID-19 and associated government policy responses. They show COVID-19-related maternal deaths worldwide (NakamuraLPereira et al. 2020), and the disproportionate loss of jobs for women for two primary reasons: the feminized nature of industries affected by lockdowns (e.g. tourism and hospitality) and the gendered norms related to childcare when schools and nurseries close coupled with restrictions to kinship care (Alon et al. 2020).

Women concurrently work in sectors most impacted by COVID lockdowns, such as hospitality and tourism and education, while shouldering the brunt of unpaid care duties which have significantly increased due to school and nursery closures (Hupkau and Petrongolo 2020), as well as paid care duties as 70% of the global health workforce. A US study showed that the gender gap in working hours between men and women has increased by 20-50% as women have responded to school and nursery closures by decreasing their working hours four to five times more than their male counterparts (Collins et al. 2020). Further studies highlight the particular difficulties in achieving work-life balance for women with children under 5 years old when schools and nurseries closed in Italy (Del Boca et al. 2020); among dual-income heterosexual parents in Australia, mothers saw far greater increases in unpaid work than fathers, however an increase in paternal unpaid work lead to an overall decrease in the unpaid work time gap between men and women (Craig and Churchill 2020). Decisions to continue in paid employment or undertake the unpaid labour within homes forms part of routine household bargaining in dual parent households, affected by social gender norms, feminised sectors of the economy and gender-pay gap. If a woman is already doing more childcare, already out of work because of lockdown, or earns less, it is likely she will have absorbed the additional unpaid labour during the pandemic. The result of this is stark: in 2020, 5.4m women in USA lost jobs compared to 4.4.m men (Kurtz 2021).

Significant gendered effects of COVID-19 on mental health due to increased domestic duties, including childcare, and women’s greater levels of psychological distress have also been reported (Proto and Quintana-Domeque 2020; Ausín et al. 2020; Xue and McMunn 2020).

Analysis of men and women’s reported worries during COVID-19 showed that women were more concerned about family and loved ones, while men worried more about society and the economy highlighting gendered differences in roles and responsibilities throughout the pandemic (van der Vegt and Kleinberg 2020). A rise in intimate partner violence has also been reported globally (Usher et al. 2020).

We set out to explore the gendered effects of the COVID-19 pandemic in the UK, to understand what impact it has had on the everyday lives of women. The UK is a reasonably high performer in gender equality globally, being 21^st^ in the World Economic Forum’s 2020 Global Gender Gap report (down from 15^th^ in 2018, (Neate 2019)) and 6^th^ in Europe according to the European Institute for Gender Equality’s Index. Key challenges to gender equality in the UK include women’s limited representation in parliament (29% of ministerial positions and of members of parliament), persistent gaps in pay and full-time employment between men and women, and women’s disproportionate unpaid care burden (Barbieri et al. 2020).

Addressing gender equality in UK government policy was first initiated by Blair’s ‘New Labour’ government in 1997 (Squires and Wickham-Jones 2004). Per the 2010 Equality Act, the government has a statutory duty to give “due regard” to the impact of their policies on people with protected characteristics which include age, disability, gender reassignment, marriage and civil partnership, pregnancy and maternity, race, religion or belief, sex, and sexual orientation (UK Government 2010). Given this, policies introduced to limit the spread of COVID-19 are subject to this duty and should have been formulated so as to prevent discrimination between genders and other groups. We sought to understand the effects of these policies from a bottom-up approach, to understand the impact of COVID-19 regulation on everyday women across the UK.

In doing so, we also sought to understand the type of gendered effects that were being witnessed, beyond the descriptive direct or indirect, we recognise that the effects of policy decisions can impact women in a myriad of ways. Using a framework developed by Kabeer and Murthy (1996), we assess the effects of the UK government’s policy interventions by evaluating gender-specific constraints that affect women because they are women, gender-intensified disadvantages that affect all genders but disproportionately impact women, and gender-imposed constraints that arise due to the gendered assumptions of policy- and decision-makers. Although these types of disadvantages can be categorised, they represent the co-evolution of norms, laws and policies and are therefore both highly inter-related and mutable. This framework allows us to more clearly identify which disadvantages could be directly addressed or alleviated through policy change (Seeley, Grellier, and Barnett 2004), recognising that policy changes to address gender-imposed constraints could be achieved more easily, and help to facilitate broader normative change that results in the gender-specific and gender-intensified disadvantages women currently experience.

## Methods

Using a gender matrix methodology (Gender & Covid-19 Project Group 2020), we searched media and grey literature to identify the key issues emerging within the UK to set out initial analysis and areas for further insight. This allowed to us to design an interview guide which covered the concerns we believed to be emerging in the early stages of COVID-19 in the UK, and to identify key constituent groups of women at risk of the direct or indirect effects of COVID-19 response for further analysis. Semi-structured interviews were conducted with 64 women between April and June 2020. Women came from 5 constituent groups who were deemed to be most at risk of the downstream effects of the outbreak, as identified through the matrix analysis. This included healthcare workers; parents (particularly of small children); migrant women; pregnant women/new mothers; and those working in industries most affected by lockdown: recreation, tourism and hospitality (ONS 2020a). Recruitment occurred via social media – by sharing information about the project with requests for participants to contact us if interested in taking part; and similar requests were shared through mailing lists from established women’s organisations in the UK. Interviews were conducted by phone or skype between 20^th^ April and 5^th^ June 2020. Given pandemic working practices, informed consent was obtained verbally at the start of the call. Interviews were recorded and transcribed verbatim. Ethical approval for this study was granted by Simon Fraser University Research Ethics Board, reference number 2020s0126 / 440243, and London School of Economics and Political Science Research Ethics Committee, reference number 1096. Six months later, as the pandemic continued, we recontacted the same participants by email and asked for a second interview. 32 women participated in a follow up interview between 21^st^ October and 20^th^ November 2020.

The 64 women interviewed were ciswomen aged between 24 and 70 years old. 52 participants self-identified as white (44 British; 8 foreign nationality) with 12 identifying as women of colour of either British (6) or foreign nationality (6). Sexual preference was not identified in the study, however all but one of the co-habiting women was living with a male partner.

Framework analysis was conducted on the transcripts obtained (Ritchie and Spencer 2002). Two researchers reviewed the transcripts identifying key trends in the data and creating an iterative coding guide for the remaining transcripts. This framework was then subject to second order analysis as consideration was given to finding synergies and trends in the data. The findings below were then overlaid on Kabeer and Murthy’s analytical framework for understanding the gendered effects of policy packages, differentiating themes identified in our data set between gender-specific constraints, gender-intensified disadvantages and gender-imposed constraints (Kabeer and Murthy 1996, as cited in Kabeer and Subrahmanian 1996, 40). Gender-specific constraints are those constraints that women experience because they are women. These may include limits to the type of job a woman can do and how much a woman can earn doing such jobs. Gender-specific constraints may also limit a woman’s time to engage in activities because of their unpaid care burden. Gender-intensified disadvantages are those that are experienced by all genders but disproportionately negatively impact upon women, particularly marginalised women, due to race, single parenthood, location, and other intersecting drivers of vulnerability. Such disadvantages can also include the unequal distribution of unpaid labour between different genders and the mental load that accompanies it. Gender-imposed constraints reflect the biases of policy- and decision-makers in the provision of public goods and services like health care and social protection, and employer biases around support to remain at work after having children. These three categories are discrete but inter-related.

## Findings

### Gender-specific constraints

Gendered norms and biases across society, within our homes, workplaces and policy-making can have direct, negative impacts on women specifically because they are women. One of the key impacts identified by women interviewed were the changes to paid and unpaid labour because of the outbreak. These changes were compounded by lockdown restrictions requiring the UK population to work at home while schools and nurseries were shut (first lockdown), and compounded by the lack of kinship care due to restrictions on household mixing and the shielding requirement for elderly people. This had myriad impacts for women.

#### Paid and unpaid labour

Most of the women interviewed, and particularly those with children, described having assumed more domestic care responsibilities during lockdown: *“I do more of the childcare because he’s working a normal working day”*. For many this meant that paid employment suffered because of the additional childcare responsibilities and home-schooling because of school and nursery closure: *“I was close to breakdown because the school was sending in an influx of work, like, you have to do this, you have to do – and you can imagine I’ve got four children in three different classes… and then obviously my work, having to do all these things remotely now from the house… everything takes that much longer*.*”* The result of this was perceived poor performance in (paid) work “*And I’ve made some mistakes and I feel I’m not really aware of what my team are up to”*. Some women had created work and care schedules with their partners typically resulting in long days and exhaustion: *“…[we’re] tag teaming it, and I typically do teatime and bath routine to let my husband work, and then once the kids were in bed and dinner had happened* 舰 *then from about half-past eight I would be on my laptop until 11 o’clock, midnight some nights*.” This practice had continued for many women and their partners over the summer until schools reopened.

In response, several women requested furlough to avoid the necessary balancing act between their paid and unpaid responsibilities, once furlough regulations changed to be permitted for childcare reasons at the discretion of the employer on July 1^st^ 2020 (UK Government 2020c): “*I have been furloughed to my great relief, and more or less at my own request, to be honest, it was kind of a mutual thing because after six weeks of having the two at home, and my husband and I both working from home and on emails until midnight, I was running myself into the ground*.*”* Another woman noted, that this ability to take a pay cut to perform unpaid care duties was tied up with living costs and the need to assume the same income *“I’ve been thinking about, can I afford to go part time, in order to just [survive], which would involve me effectively having to sell my house*.”

For women like these, furlough generally came as a relief, though for some it has further reproduced traditional gender roles and presented a challenge to their identity: “*And since then [being furloughed] I’ve been the Stepford Wife, as I’ll call myself, and my husband’s working, which has been quite a shock to the system, because I didn’t even take a full year on both of my maternity leaves”* while others were concerned that furlough would lead to inevitable redundancy, both officially as they effectively tell their employer they are non-essential and in terms of their self-worth. In the face of a second lockdown in November, one furloughed retail worker told us: *“I don’t feel I’ve got a purpose, which I kind of got from going to work. So, it’s a question of just trying to find something to do*..” For many women six months on, the reopening of schools proved a godsend that allowed them to establish greater balance between both work and family: “*having children back at school has been a massive kind of change and improvement in, in our everyday life really*”.

However, discrepancies in government policy between shops and schools re-opening meant hard choices for women working in retail who were taken off furlough but still had children at home: *“They said, you know, we’re not saying, you’ve got to come back to work, but you’ll be on unpaid leave. But I was in a position where I couldn’t go back to work because I had the three children at home. And … I thought, actually, you know what this is indirect sex discrimination here, because even though they had taken everybody off [furlough] in a blanket way, the impact really was more on women, because it was women who are generally doing the majority of the childcare and who wouldn’t be able to come back*.*”*

Organisational norms have also presented challenges in workplaces which have not recognised the burden of the pandemic on all their staff, not just working mothers, and, in some cases, have increased workloads. For one woman in a management position, it has proven difficult to counter organisational requirements with what she knows to be the realities of parenthood: *“When people have been like, ‘Oh, my God, my kid’s got to self-isolate. I’m not sure how I’ll do it this week*.*’ I’m like, ‘shit, I’m not sure how we’ll do it, because we need you*.*’ Whereas the real me would be like, ‘well, that kid comes first*.*’ … I have found it hard to be the person that I want to be in these situations, because of the pressure that we’ve been under*.*”* Six months later, one mother reported that her workplace had made substantial accommodations, showing that supportive and gender-responsive workplace policies were possible: “*They introduced like a time sheet code that was just a COVID-19 code that parents and carers could use. And just said, ‘if you can’t work the hours that you’re contracted to work, don’t worry about it*.*’*”

Increases to, or shifts in, domestic labour was a theme across all interviews, demonstrating the gendered social roles which have continued to dominate despite both parents being at home. Women have absorbed the majority of the domestic load in terms of both developmental and non-developmental activities for childcare, cooking, and cleaning etc: “*we’ve moved into “boy’s tasks, girl’s tasks” is what it feels like*.”

Whilst for some this distribution of household labour was considered a practical response to the circumstances, as a result of routine gender pay gap disparities: “*it hasn’t fallen equally in my house. And we’re both aware of that, but it comes down to how much we earn and who is the breadwinner, which in turn comes down to me having children and time now to look after children*”, some women were active in the reproduction and re-entrenchment of gender norms around unpaid labour: “*there’s no way that [this] would be doable if I was still working… because that makes it so much more manageable for him. That is a much less stressful situation for him than it was when I was still working. I have no idea what furlough’s going to mean for me*”; and on household decision making “*It’s not fair to expect him to work and then have to think about what we might want to eat that week …So I’ve been doing all that stuff*”.

Nevertheless, a number of women expressed resentment at the imbalance of their situation, as one self-employed woman told us: “*we’ve been trying to split the day, but …. He was like I can’t, he was like, I just can’t, I can’t do this anymore. So, I have to backburner my business… It’s such a struggle. I’m nowhere near where I should be… it’s the feeling that because my husband’s salaried, and he works full time and that he has to take priority because he’s, you know, we would sink without that money. So, I need to step back to allow him to do his thing. But then where does that sort of leave me?*”

In some interviews, we do see men increasing their domestic roles, but these have predominantly fallen into three categories: childcare; supporting those with pregnant partners; and supermarket shopping. Many co-habiting women reported their male partners engaging in more of the childcare, however the distribution of that labour remained unequal, with women typically taking on more of the additional load, replicating nationwide data (ONS 2020b). Some women reported that the way they engaged in childcare differed from, and was more time consuming than, their husbands’, highlighting an additional mental load experienced by women with children compared to their male partners: “*my husband would be perfectly at ease with plugging them into a tablet or putting something on our projector … I sit there and go, ooh, I must follow a load of things on Instagram and find some sensory play and make some bloody chalk paints*”

Pregnant women reported partners absorbing additional work so they could rest and/or be protected from the outside world. But we do not know if this is different to how it would have been had they been pregnant in pre-pandemic period. One pregnant woman told us: “*I haven’t personally been to the supermarket since the middle of March. My husband’s taken on all of that to make sure that I’m protected*.” Several other women also reported that their male partners had taken on more of the shopping: “*The only thing is obviously my husband is doing, kind of, all the shopping and he’s going and he’s, having to, do everything*.” And more than one interviewee warned about the return to (gendered) instincts: “*I think that we’re possibly reverting a little bit to male dominance in making decisions and going out in that hunter-gatherer role*.”

### Gender-intensified disadvantages

The COVID-19 pandemic has substantially negatively impacted people of all genders. However, as this paper lays out, women are disproportionately impacted by the UK government response to COVID-19. Furthermore, not all women were impacted in the same way, and some women were significantly more affected than others.

#### Single Parents

Our data demonstrated single parents - 90% of whom are women (ONS 2019) - were some of the most severely challenged by the lockdown measures, having to provide 24/7 childcare, and simultaneously needing to work to pay the bills. This mirrors broader national trends where 1/10 single parents lost their job and 1/3 single parents were furloughed, resulting in increasing poverty amongst single headed households (Clery, Dewar, and Papoutsaki 2020). As one parent elaborated – “*I did consider asking work if I can be furloughed, because I thought it’s just going to be much easier. But because I’m already working part-time to fit around school hours, I can’t – I just can’t financially afford to take a 20% pay cut. It’s just not possible*.”

These realities went unrecognised by government response policies, as elaborated by all single parents interviewed: “*[The] implication was that you know, your partner or somebody in your bubble should be doing that. But as a single parent, that isn’t an option for me. I feel like I’ve had to repeatedly perform single motherhood and keep making this, this this point that the realities of parenting during this situation are just not being recognized that way*.*”*

The clash between financial concerns and parenting concerns was acute amongst all single parents interviewed. This led to further gender-intensified disadvantage: the mental health and emotional burden that many women suffered from during lockdown. Over the six-month period between interviews, single parenthood had been exhausting for women at home and in their attempts to gain support from employers: “*I’ve been exhausted, and I know that everyone around me is exhausted as well. And so, you know that everyone’s in the boat in the same boat, and just experiencing it in different kinds of ways. But yeah, I think I’ve literally just gone into survival mode and just trying to get by really*.”

#### Mental health

The mental health burdens varied between women interviewed but were substantial across all demographics. Almost every woman we interviewed referred to some sort of stress or tension. Some were expressly worried about the virus: “*[I’m] waking up and finding it really hard to breathe, and being worried is this COVID or whatever, but actually realizing no this is actually just panic and anxiety in my chest*”. Some felt guilty or concerned about the impact of the lockdown on their children or their dual burden of paid and unpaid work which have merged because of the lockdown: “*I worry about my daughter mostly, because she I think is just missing out on an awful lot at a crucial preschool development stage, you know*”; “*because I’m working, he’ll just be watching stuff on his iPad, and so I feel bad about that*”. Others are overwhelmed by the sheer load of the paid an unpaid care that they are undertaking and feel as if they have reached their bandwidth for being able to cope. “*I just can’t cope, to be honest. Just working and managing children at the same time, indefinitely, it’s so hard. You can’t do anything*.”

These mental health concerns were compounded by the lack of usual support networks which women might utilise as mental and emotional support in their dual roles. “*I’m also quite aware, especially with a couple of my friends, where I would ring them up and have a coffee and a chat, I’m aware that they have caring responsibilities at the moment*”. This also affected care: *“being a single parent, I’m in a bubble with my parents, but that makes it much more difficult for us to see friends which I think certainly now, you know, we can only meet up outside with other people in a, you know, in a park or outside walk, which during the winter, it’s just not as easy, pouring down, it’s cold*.*”* A number of women reported greater levels of isolation during lockdown and relief when lockdowns ended: “*It’s been a lot better since lockdown eased. I think I was really feeling the isolation during lockdown*.*”*

Six months on, women reported high levels of exhaustion and that their mental health had been up and down, with the oncoming winter and additional lockdowns being a particular concern: “*I’m having this malaise where it’s just like, my energy was so like, knowing that another lockdown was coming, knowing that winter was coming, … what was coming around the corner next, you just were like, I can’t, can’t do more of this*.”.

One woman’s experience echoed earlier interviews regarding a loss of sense of self: “*It did get to a point when I kind of said actually, you know what, I am going back to work because I didn’t feel I just didn’t feel as though I was being particularly valued by anybody*.*… None of the children ever thanked me for my particularly enriching home-schooling lesson that I’d helped facilitate for them. And, you know, if they were going to be grumpy about it, it was generally with me. And it was incredibly intense, all of us being at home all of the time. And I think I just felt like I was facilitating everybody else’s life. And mine had just ground to a halt, it was almost like, going back to the 1950s*.”

Nevertheless, many women highlighted the support they received from their network of friends and support networks: “*One of the things that I think is really great for me, is that the women in my life have an amazing kind of network constantly asking each other about mental health. And I think we’re trained to do that from the baby groups, the teachers are always saying, ‘don’t forget, check in on each other and make sure everyone’s alright’”*.

#### Pregnant women

The health system has been distorted as COVID-19 care and patients have been prioritised within hospitals, and as efforts have been made to reduce interaction between individuals (Charlesworth 2020). Many non-essential services have ceased or have changed to online provision. As women are more likely to interact with primary health providers than men (Wang et al. 2013), changes to healthcare provisions have a gendered effect. Moreover, our participants included pregnant women, and health system distortions had a considerable impact on them (anonymous, forthcoming).

As one woman stated, ante-natal provision was stalled because of the outbreak: “*My midwife has phoned check[ed] in on me and – but there is only so much you can do over the phone*”. Those who were able to access care for check-ups noted the differences in how services were provided: “*I went into the hospital and the midwives wouldn’t put the devices on my belly, I had to fit them myself while they stood at a distance*”. Some women expressed concern about the quality of care: “*it does make you feel a bit more apprehensive I think, because obviously, you’re not getting the same sort of care that you normally get, like urine samples and I haven’t taken my blood pressure … you’re meant to have that done every four weeks*.”

Having to attend ante-natal care alone, as per the UK COVID-19 regulation which restricted birth partners in efforts to reduce social contacts, was a concern to participants. Notably, this decision was reversed in December 2020 (Bremner 2020). “*My 20 week scan I had to do alone, and so if there had been any issues at that point I would’ve found out by myself. That was a bit scary*”. Yet the lack of partner’s involvement was not just for emotional support during antenatal scanning: some expressed concern as to how partner’s absence may affect the quality of care they receive: “*I am … very concerned about my ability to advocate for my own care. I know that, as it stands, if things are very straightforward my husband will be able to be there for the majority of that period of time, but there will definitely be a time after the baby’s born when he can’t be present. And I am concerned that I’d really be relying on him to know what I want to be done and how I want that, so I am very concerned about that*.”

Women also expressed concern that men were missing out on the experience of parenthood, highlighting the breadth of mental burdens experienced by pregnant women during the pandemic, who worried not only about themselves and their babies, but also their partners: “*I just think that the effect and the impact on him is probably bigger than even on me, because he isn’t able to participate in anything that’s really important to both of us*”.

This concern of a mismatch between expectations of pregnancy and the reality facing women is also important. Women had imagined that they would be able prepare for becoming a parent - to try out prams, attend antenatal classes, introduce their baby to their family when born etc – each part of the ritual of pregnancy and new motherhood (Afflerback et al. 2014) which they were no longer able to enjoy: “*And sad, I think, more than anything about missing the fun things that are supposed to happen when you’re pregnant. Like a baby shower, and going shopping for a stroller, and stuff like that. Such minor issues compared to what most people are dealing with, but it’s just you look forward to these things*”.

This also extended to social networks which could not be utilised, but which new mothers are reliant on: “*being a first-time mum it’s obviously quite daunting in this, so you need a lot of reassurance, not, not being able to have, say, for example, my mom here and things like that, you’re doing everything over the phone”; “there was all that stuff about like, oh, Mums shouldn’t be gossiping at the school gates as if like, oh, we’re all so silly, and we’re gossiping. But actually, what if that’s the only person that you see all day? What if that’s the only other adult that you get to speak to all day?*”

### Gender-imposed constraints

We sought to explore why it was that these gendered-specific constraints and gender-intensified disadvantages had emerged in experiencing lockdown in the UK. Further analysis of the data highlighted gender-imposed constraints, the result of government decision-making.

#### Representation in government

Several respondents noted the lack of representation in government decision-making and in communication to the government, and attributed the lack of recognition of gendered effects of COVID-19 related policy to the fact that there weren’t women ‘in the room where it happened’. “*Almost every night I told my husband, oh, no women on the screen [for the government press conference], oh, no women on the screen. I mean, it doesn’t mean that men are bad, obviously, but I think, again, if we only show men in power, what does it say about women?*”. The concern amongst participants was that this contributed to unconscious bias whereby women’s concerns were not considered in the development of COVID-19 response policies. Two highlighted: *“you’ve got somebody that’s white male and middle-aged telling the nation, predominantly – well, predominantly white male middle-aged - what we should and shouldn’t be doing”;* “*I just don’t think they think about women because they haven’t got enough women in them to, you know, they don’t have that lived experience. So, it’s just it’s always an afterthought, isn’t it?”*

For many this lack of women’s representation contrasted with the role women were playing on the frontline of the response, highlighting the mismatch between the risks and additional work that these women were doing and their lack of meaningful recognition by the government. “*The majority of NHS staff seem to be women and they are considered our heroes, but they don’t actually get funded*”; “*You know, don’t go ‘oh love,’ because it’s a bit patronising isn’t it, ah, you lovely nurse, you lovely carer, you’re going in there to look after the old people… and they’re potentially going to die on £6 an hour without a mask? Like they don’t give a shit about your clapping [Clap for Carers became a weekly activity in the UK during the first months of COVID-19]*”

However, these gender-imposed constraints also raised questions about how men and women are considered differently in public life and within families. As one respondent posed – “*why are we not witnessing a conversation about men doing more, rather than analysing the suffering of women*?” This applies from the top down: “*no one’s asked Boris [Johnson] what his children are doing or – so I still feel that there’s very ingrained … roles within society that are very much female rather than male*”.

While some women felt the government had done their best in a difficult and ever-changing situation, several women felt there had been little change in the government’s approach to considering and supporting women throughout the pandemic, despite increasing research and public awareness: “*I still think that it’s expecting women to pick up the strain… the Conservative government don’t care about women, they don’t care. They don’t care about a lot of people, but particularly, they don’t care about women. And I think they’ve got quite an old-fashioned view, outdated view, that women will just pick up the childcare, the women will pick up the care of elderly parents, they will shoulder the burden of whatever policies are put in place, or decisions that are made, they will just get on with it. And these, I haven’t seen anything that makes it easier for women - all I’ve seen the things that make it more difficult for women*”. There was also concern for the longer-term impacts of this perceived gender blindness: “*It’s always bolted on and I honestly think that the damage this is going to do for you know, women in the workplace. This again, it’s going to be really far reaching, it’s going to go on for years. I think we’ll have to claw it back*.”

A number of women living in the north of England expressed frustration at the government’s perceived neglect of the lowest socio-economic and minority ethnic groups, and the intersectional impacts of policy interventions that intensified gender-imposed constraints: “*I think the glaring inequalities have been in class, I think more than more than women. I think this has shown the glaring inequalities in class. And you can see that by the number of number of BAME people being sick and the link to an obviously not it’s not just a link because obviously there’s other issues around BAME getting COVID. But you know, there’s a clear link between poverty and having to work and work in services that might be vulnerable to COVID. So no, I don’t think women have been considered and obviously, women have been fucked because of childcare and if you can’t afford childcare, then how can you go to work?*”

## Discussion

Our data show a range of indirect effects on women of COVID-19 and associated non-pharmaceutical interventions instigated by the UK government. Whilst lockdown can be justified from an epidemiological perspective to reduce the circulation of the virus, due consideration needs to be given to the secondary effects of this public health intervention, and how these effects are experienced unequally across society. As shown here, the effects of COVID-19 and associated government interventions to minimise disease transmission have significant gendered constraints, impacts which are intensified by gender, and those which are imposed by macro and micro gendered norms. In line with the Public Sector Equality Duty (PSED) under the Equality Act 2010, government representatives insisted that COVID-19 policies were subject to equality assessments (Pyper 2020, 27–29), though these assessments went largely unpublished.^1^ Without real-time publication of these assessments, and in light of our findings and those of others, questions remain as to the rigour of government analyses: the task of assessing government policy lies with civil society and academics instead (Conley 2012). We write this paper for policymakers to understand the downstream effects of their COVID-19 decision making, and to consider these impacts if and when they are forced to implement additional lockdowns (as we write during the UK’s third national lockdown).

Our data on women’s additional unpaid labour mirrors those of other studies quantifying the additional toll on women’s workload during lockdown (ONS 2020b), and the impact of COVID on women’s labour force participation because of the macro and micro economic decisions. These include the effects of sector-wide lockdowns in heavily feminised industries like hospitality, tourism and recreation; of gender-pay gaps and the need for the higher [male] earner to continue to work (notably at a time where the UK government suspended gender pay gap reporting for employers (UK Government 2020b)); or the gendered division of labour within households such that women have borne the brunt of the additional childcare (Adams-Prassl et al. 2020). Our research adds women’s voices to these statistics, bringing nuance to the numbers. As one woman summarised: “*you’ve got to be a superhero, you’ve got to look after your kids, you’ve got to manage the house, you’ve got to keep your husband out of the way, and get on with the other life that you normally do, which is working as well, and somehow fit it in*.”

It also allows insight into how divisions in labour have emerged, how women understand their paid/unpaid work in relationship to their partners within domestic ideologies of motherhood (Dyck 1990); with some women feeling that their careers are secondary, and that they have to give up work, reduce their hours, or work anti-social hours to facilitate their husband’s routine workday. Indeed, as is well established, women’s ability to engage in paid work is entirely dependent on their unpaid care responsibilities, and vice versa, thus both areas need to be considered by government policy (Perrons 2005). In many ways our data shows COVID-19 as an amplification of the motherhood penalty (Correll, Benard, and Paik 2007), but this penalty has been exacerbated because of the pandemic circumstances, rather than active decision making by a couple. Indeed, it appeared that the decision-making was tacit, reflecting gendered norms and implied household bargaining within families interviewed. Even those women who were frustrated by the gender constraints and intensification of these, remained resigned to them, reinforcing gendered power differentials within UK households and society.

The demands of paid labour and increases in unpaid labour have taken a toll on women: whether in a loss of identity having to give up their work, or with more acute mental health concerns as a combined result of the risks of the virus, worry about their children, their financial concerns (or a combination of all of these), and a sense of being at their emotional bandwidth, unable to take more stresses and trying to juggle these competing demands. The responsibility for keeping life going, and assuming the emotional burden of the family, being the sounding board for upset and bored children also appeared heavily feminised. As one mother described: “*I hold it all – I do hold it all together*.*”*

These impacts were particularly intensified amongst single mothers who were suffering the same impacts of having to juggle paid and unpaid work, and the associated anxiety, but didn’t have the same physical or emotional support as dual parent households. Whilst single parents may be accustomed to navigate the additional challenges posed, through a range of social networks and established coping strategies (Defrain and Eirick 1981), the pandemic has cut off many sources of support to them through stay at home orders and the inability to draw on kinship care. This might be caring arrangements with family, friends, or formal childcare, and their ability to interact with other adults for their emotional well-being. Furthermore, full-time caring responsibilities have placed some single women in even more precarious financial positions, well documented to led to greater anxiety (Stack and Meredith 2018), and indeed exacerbated the risk for some single parents of falling into poverty (Cain 2016).

Pregnant women were concerned about changes to access in maternity care, and how this brought safety concerns to their pregnancy. Whilst the NHS maintains that safe access to maternity care was not jeopardised by the pandemic (Rimmer and Wattar 2020), women’s perceptions of being able to access such services may have altered their interaction with maternity care, which caused added anxiety during pregnancy. Further concerns have been raised about the impact of changes to delivery options and women’s decision making in this process on how women understand and experience pregnancy, which in turn can lead to heightened risk of post-partum depression (Bell and Andersson 2016). Beyond the health concerns, women simultaneously grieved the rites of passage of pregnancy that they missed out on because of lockdown. A 2017 study in Yorkshire, UK highlighted men’s psychological distress during their partner’s perinatal period and feelings of exclusion regarding maternity services (Darwin et al. 2017). Limits on the presence of birth partners is likely to have exacerbated these issues for expectant fathers.

We can start to understand how these gendered impacts have come about – in a top-down manner through government actions and policy creation which failed to recognise the gendered nature of lockdown, and from the bottom up through the reproducing of gendered expectations and social understanding within households, collectively imposing gendered constraints on women. At the macro level, we assume gendered norms held by government representatives has led to gender blindness in policy-making and implementation^2^. Although the UK government is required under the Equality Act (2010) to ensure all policy produced undergoes an impact assessment, the government have refused to publish their assessments. At the same time as the COVID-19 policies were being developed, the UK government halted the requirement for organisations to report gender pay gap data. This is reflective of the broader position given to gender issues within the current administration, which has not only reduced capacity and funding to the government equalities office within the cabinet office, but has also merged the position of the Secretary of State for Women and Equalities with the function of the Minister of International Trade (UK Government 2020a), the latter being of much more importance during Brexit negotiations, and thus time commitment to government activities. A review of four European states’ economic and social policies during COVID-19, found UK policies to be lacking compared to those in Norway (whose furlough scheme was more generous for low-income earners while the UK’s income replacement rate remained the same), Germany (which gave additional financial support to families with children) and Italy (which, along with Norway, increased parental leave for new parents). Cook and Grimshaw (2020) highlighted the gendered assumptions of UK government policies which “assume a normative (male) worker and leave the gendered division of domestic labour unchallenged”. This also needs to be understood within the broader context of Brexit, given that most progress for gender equality and mainstreaming in the UK was driven by requirements under EU employment, maternity and childcare directives (Fagan and Rubery 2017). Whilst the government has assured constituents that equality rights will be maintained once the UK has left the EU, there are no legislative commitments (EIGE 2020).

At the micro level, women were also reproducing these gender-imposed constraints in the way in which they discussed the gender-specific impacts experienced, such as the distribution of and decision-making concerning paid and unpaid labour during lockdown. As such, it appears that women’s participation in public and private activities was determined by longstanding gender norms at macro and micro levels, and as defined in relation to how men interacted with these issues, rather than as a prioritisation of their own needs. This is an important finding as it allows reflection in the production of these gender-specific constraints, gender-intensified disadvantages and gender-imposed constraints, which arose as a result of a combination of staunch gender norms across households, society and government.

Similar gender norms appear throughout our data, demonstrating perpetuating gender norms within the micro level of the section of UK society which we interviewed. Whilst some women discussed the distribution of paid and unpaid labour, and the emotional toll in practical terms, others recognised the distinct gender-imposed constraints, and yet appeared to be unable to alter these amid the ‘tyranny of the urgent’. These gendered norms, and the differential impacts that women have experienced in the UK during 2020 must be taken seriously and considered during after-action reviews to ensure steps can be taken to minimise further unequal impacts on women as a result of COVID-19 related disease interventions.

Moreover, we believe it is important to differentiate the gendered effects of the pandemic, as we have with gender-specific constraints, gender-intensified disadvantages and gender-imposed constraints so as to be able to understand the agency of different actors for change. For example, whilst gender-specific constraints may be predominantly at the micro level within household decision making for care based on entrenched social norms, there are government policy developments which could mitigate some inequalities, including efforts to reduce the gender pay gap, or concerted efforts to reduce the differentiation between feminised and masculine sectors of employment, given that pandemics can affect different sectors owing to social distancing. Moreover, understanding gender-intensified disadvantages allows for consideration of the broader socio-economic determinants of health, and of government policy. This allows efforts for change to not be limited to women, but to understand the breadth of the impact of COVID-19, and the inequalities experienced across societies. Indeed, the increasing engagement of men in childcare and domestic duties, whilst still less than women during the pandemic, highlights how the realm of unpaid labour might nowadays be considered more of a ‘gender-intensified disadvantage’ than a ‘gender-specific constraint’ that can be further facilitated through government policies that support men’s engagement as carer during the perinatal period and during the early years of a child’s life (see Lewis and Campbell 2007 for a review of UK work/family balance policies, 1997-2005).

Gender-imposed constraints are also mutable, through efforts for greater representation in decision making, but also through the inclusion of gender advisors and analysis in decision making.

These gendered impacts are vital to understand and address in policymaking. There is a risk that whilst gender equality was already slipping in the UK prior to the COVID-19 pandemic, that the impacts experienced by women during 2020 can have long-lasting effects, particularly the gender-imposed constraints and their being forced out of the labour market. Coupled with an unsupported childcare sector (Lewis and West 2017), a significant reduction in female participation in the labour force and an increase in the gender pay gap could last for a number of years (Alon et al. 2020).

This analysis has focused on the experiences of women from across the UK and in relation to Westminster interventions. Westminster’s commitment to the Equality Act 2010 has not been as strong as that in the three devolved administrations of Wales, Scotland and Northern Ireland (Hankivsky, de Merich, and Christoffersen 2019) which is also reflected in the Equality and Human Rights Committee’s suspension of PSED compliance activities in England but not in Wales or Scotland (EHRC 2020). Further studies on the differing responses by devolved administrations and their impacts could highlight areas for improvements with the Westminster policy-making process to ensure that all women across the UK are better supported during the remainder of the pandemic and through future policy initiatives.

## Data Availability

Data is available for review upon request.

## Footnotes

1 One exception is the Department of Health and Social Care’s public sector duty impact assessment of the Coronavirus Act 2020, published on July 28, 2020 – four months after the initial publication of the Bill. Yet, the assessment does not review the entirety of the government response, only those issues related to the department itself (DHSC 2020).

2 We have not interviewed government representatives to understand their understanding of gender, but we make this assumption due to the lack of interaction we have seen on these issues, compounded with the comments made by senior government officials when asked about gendered issues within press conferences and media publications.

